# Association of Right Ventricular Dilation and Dysfunction on Echocardiogram with In-Hospital Mortality Among Patients Hospitalized with COVID-19 Compared with Other Acute Respiratory Illness

**DOI:** 10.1101/2022.06.29.22277073

**Authors:** Kaiwen Sun, Emily Cedarbaum, Christopher Hill, Sithu Win, Nisha I. Parikh, Priscilla Y. Hsue, Matthew S. Durstenfeld

**Author notes:** Corresponding author: Kaiwen Sun, MD, Address: 1001 Potrero Ave, 5G, Division of Cardiology at Zuckerberg San Francisco General, San Francisco, CA 94110. Disclosures: The authors have no disclosures relevant to this paper.

## Abstract

**Background:** Although right ventricular (RV) dysfunction is associated with mortality in acute COVID-19, the role of RV dilation is uncertain. The prognostic significance of RV dilation and dysfunction among hospitalized patients with acute COVID-19 compared to other respiratory illnesses.

**Methods:** We conducted a retrospective cohort study to examine 225 consecutive adults admitted for acute COVID-19 and 6,150 control adults admitted for influenza, pneumonia or ARDS who had a clinical echocardiogram performed. We used logistic regression models to assess associations between RV parameters and in-hospital mortality adjusted for confounders.

**Results:** Among those with COVID-19, 48/225 (21.3%) died during the index hospitalization compared to 727/6150 (11.8%) with other respiratory illness (p=0.001). Independent of COVID-19, mild and moderate to severe RV dilation were associated with 1.4 and 2.0 times higher risk of inpatient mortality, respectively (95%CI 1.17 to 1.69; p=0.0003; 95%CI 1.62 to 2.47; p<0.0001, respectively). Similarly, mild and moderate RV dysfunction were associated with 1.4 and 1.7 times higher risk of inpatient mortality (95%CI 1.10 to 1.77; p=0.007; 95%CI 1.17 to 2.42; p=0.005, respectively). Relative to normal RV size and non-COVID-19 acute respiratory illness, mild and moderate RV dilation were associated with 1.4 times and 2.0 times higher risk among those without COVID-19 and 1.9 times higher and 3.0 times higher risk among those with COVID-19, with similar findings for RV dysfunction. Having both RV dilation and dysfunction or RV dilation alone were associated with 1.7 times higher risk while RV dysfunction alone was associated with 1.4 times higher risk compared to normal RV size and function.

**Conclusions:** RV dilation and dysfunction are associated with increased risk of inpatient mortality among those with COVID-19 and other respiratory illnesses. Abnormal RV findings may identify those at higher risk of short-term mortality from acute respiratory illness including COVID-19 beyond other risk markers.

## INTRODUCTION

Although acute SARS-CoV-2 infection is mainly characterized by pulmonary manifestations, cardiac effects may occur among hospitalized individuals with COVID-19. Up to 36% of hospitalized patients experience myocardial injury^1^ and elevated cardiac biomarkers portend worse outcomes.^1-3^

Right ventricular (RV) abnormalities are more commonly seen than left ventricle (LV) abnormalities in acute COVID-19.^4,5^ The adverse effect of COVID-19 on the RV is unsurprising as prior studies revealed RV dilation or dysfunction are the most common echocardiographic findings in those with acute respiratory distress syndrome (ARDS)^6,7^ and RV dysfunction is associated with mortality in ARDS.^8^ While substantial evidence suggests that RV dysfunction is associated with in-hospital mortality in acute COVID-19, ^9-11^ the prognostic implications of RV dilation alone are less clear. While some studies have found an association between RV dilation and mortality^12,13^, others have not.^14-16^ Similarly, little is known about the prevalence and prognostic significance of RV dilation among hospitalized patients with acute COVID-19 compared to other respiratory illnesses.

Therefore, we studied the association of RV dilation and RV dysfunction with mortality among adults hospitalized with COVID-19 compared to adults hospitalized with other respiratory illnesses (influenza, pneumonia, and ARDS).

## METHODS

### Study population

We used a retrospective cohort study design to examine 225 consecutive adults (over 18 years old) admitted for acute COVID-19 between March 2020 and February 2021 at three UCSF Health hospitals and Zuckerberg San Francisco General Hospital. Also included in the study were 6,150 control hospitalizations for influenza, bacterial pneumonia or acute respiratory distress syndrome, including both historical controls from 2019 and contemporary controls with negative COVID-19 PCR testing on admission from April 2020-2021. All patients had a clinically-ordered transthoracic echocardiogram performed during their hospitalization. Patients were excluded from the COVID-19 group if their indication for admission was unrelated to symptomatic COVID-19, such as admission for elective procedures in those with a history of positive COVID-19 test.

### Clinical characteristics

Patient demographics and clinical characteristics including past medical history, vital signs, admission labs, admission electrocardiogram, admission chest imaging, medications, echocardiographic report, and discharge disposition and diagnoses, were extracted from the electronic medical record (EMR) by the UCSF Clinical and Translational Science Institute. Through manual chart review, we verified all cases of COVID-19, all deaths among those with COVID-19, and clinical outcomes among those with COVID-19. Data unable to be extracted were manually retrieved through chart review and documented in REDCap. The main predictor variables of interest were the presence of RV dilation or dysfunction in those with and without COVID-19. The primary outcome was in-hospital mortality.

### Echocardiographic RV Parameters

For those who had more than one echocardiogram performed, we included only the first echocardiogram after the first positive SARS-CoV-2 test or admission to the hospital. Visual assessment for RV dilation and dysfunction was made by the initial echocardiographer who interpreted the clinical echocardiogram during hospitalization, and findings were extracted from the clinical echocardiogram report. Subsequently, in the subset with COVID-19, measurements of the RV were performed independently by two blinded reviewers. The kappa statistic and intraclass correlation (ICC) were computed to measure inter-rater reliability for categorical and continuous variables, respectively.

In addition to the qualitative assessment reported by the reading echocardiographer, we considered three measures of RV function including tricuspid annulus planar systolic excursion (TAPSE), right ventricular systolic excursion velocity (s’), and RV fractional area change (FAC). In accordance with ASE Guidelines, we defined abnormal RV parameters as RV basal diameter > 42 mm, mid diameter >35 mm, TAPSE <1.7cm, RV s’ <9.5cm/s, and RV FAC<35%.^17^ In a subset, we also performed LV strain measurements. RV strain measurements were not feasible on a very high proportion of these clinically obtained studies, so we were unable to measure strain in a meaningful sample.

### Statistical Analyses

We constructed logistic regression models to estimate associations between echocardiographic parameters and in-hospital mortality among patients hospitalized with COVID-19 and control patients hospitalized with other respiratory illness (influenza, pneumonia, ARDS). We only included the index hospitalization for individuals with COVID-19, but we allowed control participants with multiple eligible admissions with echocardiograms to be included each time they met the eligibility criteria. We accounted for clustering by patient in estimating the standard errors. We adjusted for potential confounders including age, sex, body mass index, race/ethnicity, past medical history, admission estimated glomerular filtration rate and hemoglobin, and for models that included echocardiograms, we included mechanical ventilation at the time of the echocardiogram. We estimated adjusted relative risks and adjusted risk differences using post-estimation. Using the same models, we estimated the area under the receiver operating curves incorporating echocardiographic parameters into the model. When possible, we treated RV dilation and RV dysfunction as three-level categorical variables (normal, borderline to less than moderately reduced “mild”, moderately reduced or worse “moderate to severe”), but for analyses where there were not enough patients with COVID-19 in each group such as when considering RV dilation and dysfunction together, we collapsed it into a binary variable (normal, abnormal). For continuous variables (age, BMI, LVEF, peak tricuspid gradient, estimated glomerular filtration rate, hemoglobin) we used restricted cubic splines due to departures from linear relationships.

Because the independent censoring assumption does not hold when comparing individuals discharged alive with individuals who remain hospitalized, we used Fine and Gray competing-risks regression models and robust standard errors to estimate the effect of RV dilation on mortality treating discharge alive as a competing risk and report the sub-distribution hazard ratios.^18^ For time-to event analyses, we started follow-up time at hospital admission to prevent immortal time bias from admission to echocardiogram. We assumed that variables were missing completely at random and therefore we used complete case analyses. Because most patients who received ICU care were already in the intensive care unit when the echocardiogram was performed, we did not examine whether echocardiographic parameters were associated with ICU transfer and we excluded those already intubated in our model for mechanical ventilation. Given our focus on the RV, we conducted sensitivity analyses excluding those with a history of pulmonary arterial hypertension.

Statistical analyses were performed using STATA 17.1. The UCSF IRB approved this study and granted a waiver of consent.

## RESULTS

### Patient Characteristics and Hospital Course

We included 225 consecutive adults hospitalized with COVID-19 from March 2020-February 2021 and 6,150 controls hospitalized with non-COVID acute respiratory illness who underwent clinical echocardiography. O the controls, 2,412 (39%) were admitted prior to March 1, 2020 (the approximate start of the pandemic in San Francisco), and 3,738 (61%) were contemporary controls admitted after the start of the pandemic. The mean age was 63 years old in both groups (p=0.99, Table 1). Compared to the control population, the acute COVID-19 group had a higher proportion of males (66% vs 54%, p<0.001) and more identified as Hispanic (38% vs 15%, p < 0.001). Those with acute COVID-19 had a higher body mass index (29.4 vs 27.9, p = 0.008). However, history of heart failure, atrial fibrillation, cancer, asthma, chronic obstructive pulmonary disease, and pulmonary embolism (PE), and pulmonary hypertension were more prevalent in the control group.

**Table 1.**
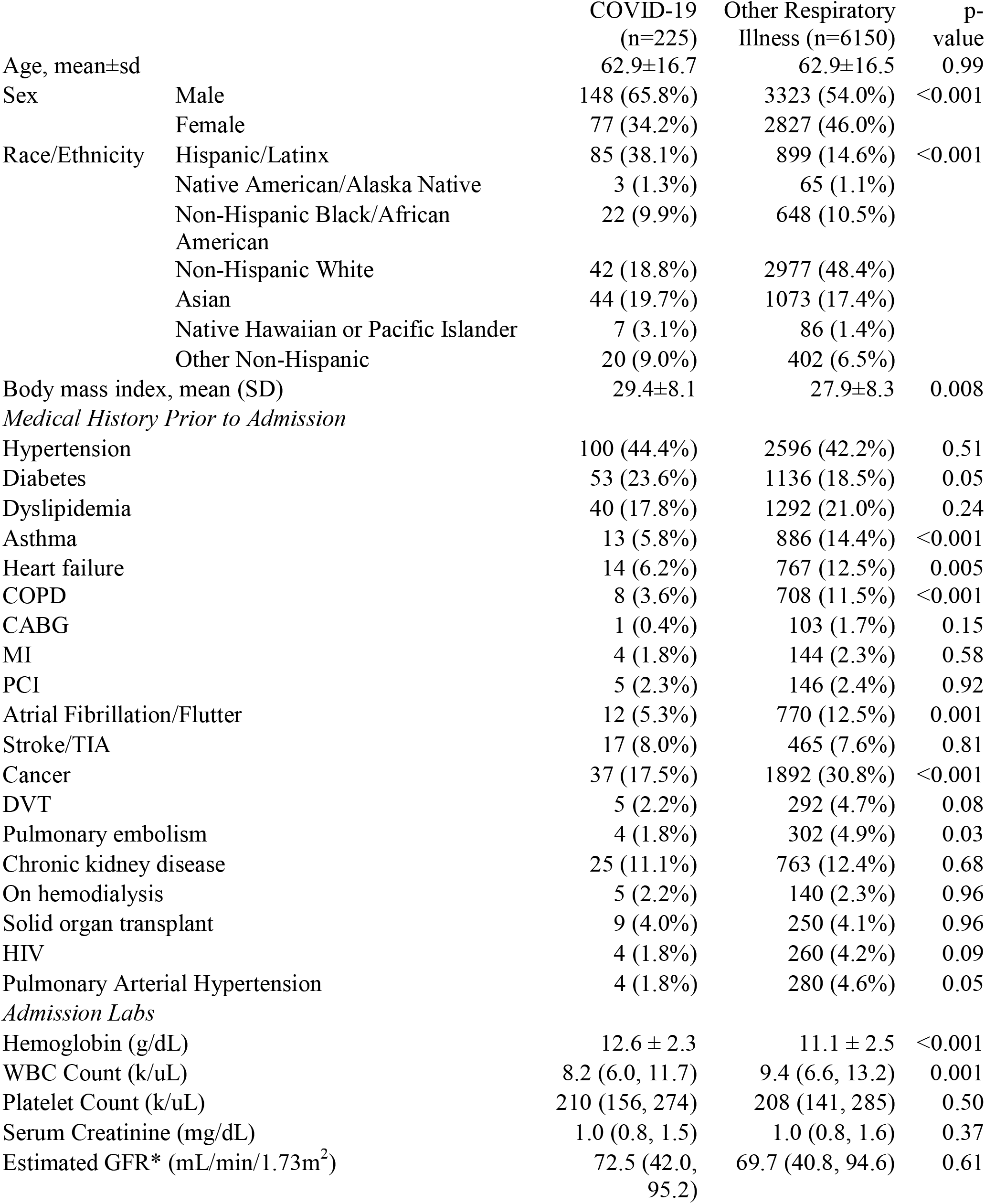
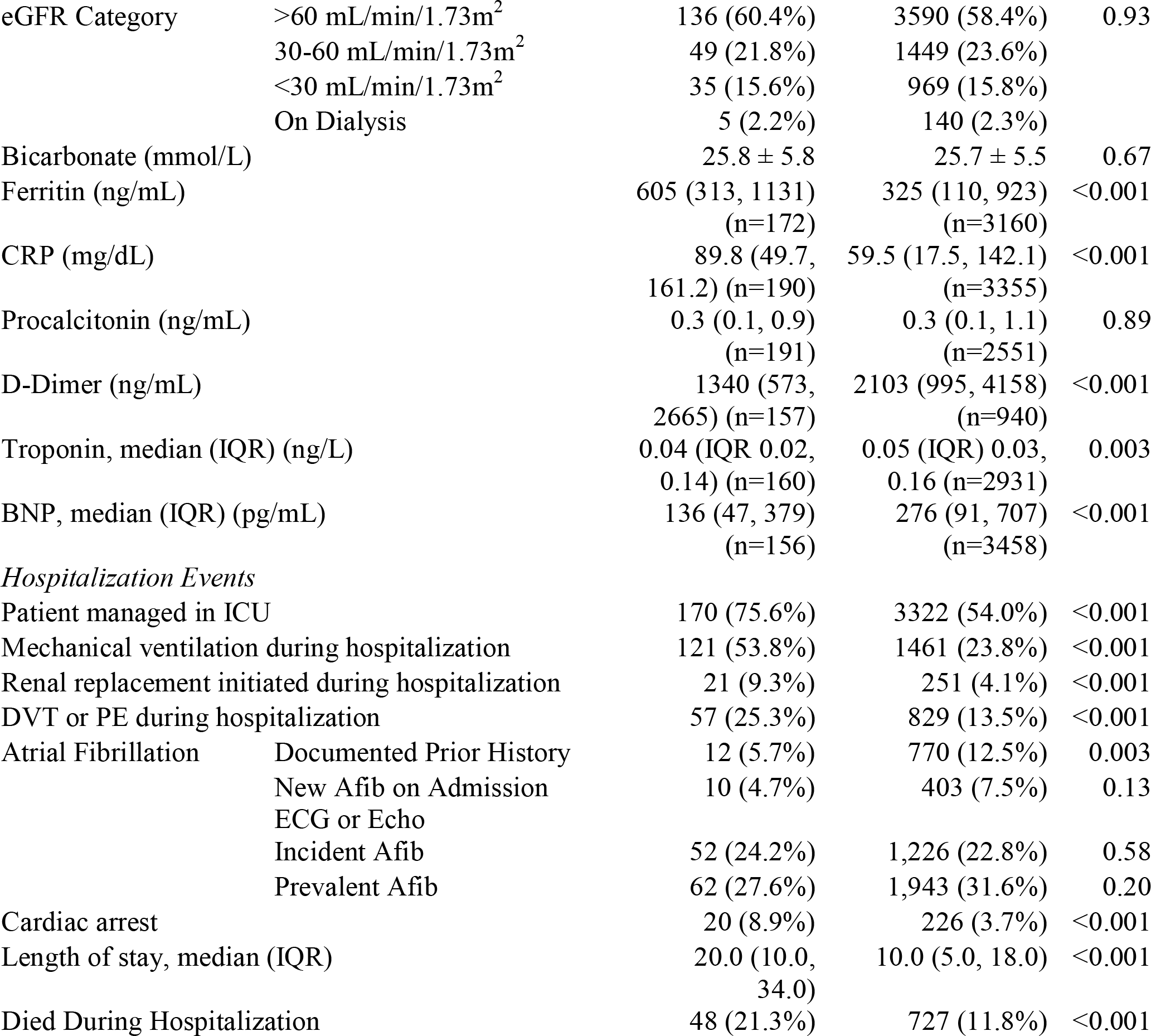
Baseline Characteristics of Hospitalized Individuals who Underwent Clinical Inpatient Transthoracic Echocardiography with COVID-19 compared to Other Respiratory Illness A comparison of patient demographics and hospital events between the COVID-19 and control group of individuals hospitalized for other respiratory illness. Other than where noted (specific labs, for example) fewer than 1% were missing among those with or without COVID-19. Estimated glomerular filtration rate was calculated using the CKD-EPI equations without adjustment for Black Race.

In comparison with controls, those with COVID-19 had higher hemoglobin and inflammatory markers and lower WBC, BNP, and D-Dimer levels. The COVID-19 group more often required ICU care (75% vs 54%, p<0.001), mechanical ventilation (p<0.001), and initiation of renal replacement therapy (p=0.002). DVT or PE was more frequently diagnosed in the acute COVID-19 population (25% vs 14%, p<0.001). Median length of stay among those hospitalized with COVID-19 was 10 days longer than their control counterpart (20 vs 10, p<0.001).

Among those with COVID-19, 48/225 (21.3%) died during the index hospitalization compared to 727/6150 (11.8%) with other respiratory illness (p=0.001). Adjusted for age, sex, body mass index, race/ethnicity, past medical history, and admission hemoglobin and estimated glomerular filtration rate, adults hospitalized with COVID-19 had 1.54 times higher relative risk of inpatient mortality compared to adults hospitalized for influenza, pneumonia, or ARDS (95%CI 1.06 to 2.24; p=0.02) or an absolute risk difference of 5.2% (95%CI 0.1 to 10.2%; p=0.047).

### Echocardiographic Characteristics

Patients hospitalized with acute COVID-19 were more likely to be intubated at the time of echo compared to the control group (31% vs 9%, p<0.001, Table 2). The most common echo abnormality in both groups was RV dilation (22%, p=1.00). Among those with COVID-19 and qualitative RV size reported (n=211), 33 (15.6%) had mild RV dilation, and 14 (6.6%) had moderate to severe RV dilation. Among the controls with RV size reported (n=5,659), 803 (14.2%) had mild RV dilation, and 446 (7.9%) had moderate to severe RV dilation. In those with COVID-19 and qualitative RV function reported (n=214), 187 (87.4%) had normal RV function, 20 (9.4%) had mild RV dysfunction, and 7 (6.6%) had more than mild RV dysfunction. Among controls with reported RV function (n=5,420), 440 (8.1%) had borderline or mild RV dysfunction, and 141 (2.6%) had greater than mild RV dysfunction. The distribution of RV size and function were not significantly different by COVID-19 status (p=0.70 and p=0.67, respectively).

**Table 2.** Echocardiographic Parameters Comparison of echo features between the COVID-19 and control groups with n listed if missing >1%. Abbreviations: TAPSE= Tricuspid Annulus Planar Systolic Excursion.

| Variable |  | COVID-19<br>(n=225) | Other Respiratory<br>Illness (n=6150) | p-value |
| --- | --- | --- | --- | --- |
| Days from Admission to echo, median (IQR) |  | 2 (1, 4) | 2 (1, 3) | 1.00 |
| Intubated/mechanical ventilation at echo |  | 70 (31.1%) | 573 (9.3%) | <0.001 |
| LV ejection fraction, mean (SD) |  | 60.4 (12.7) | 59.9 (13.8) | 0.45 |
| Reduced LV function (LVEF <50%) |  | 27 (14.2%) | 957 (17.5%) | 0.24 |
| LV Global Longitudinal Strain, mean±SD |  | 18.2±4.0<br>(n=30) | 17.4±4.1<br>(n=406) | 0.31 |
| RV volume | Normal | 164 (77.7%) | 4,410 (71.7%) | 0.97 |
|  | Borderline | 9 (4.3%) | 181 (2.9%) |  |
|  | Mildly reduced | 19 (9.0%) | 487 (7.9%) |  |
|  | Mild to moderately reduced | 5 (2.4%) | 135 (2.2%) |  |
|  | Moderately reduced | 9 (4.3%) | 244 (4.0%) |  |
|  | Mod. to severely reduced | 1 (0.5%) | 53 (0.9%) |  |
|  | Severely reduced | 4 (1.9%) | 149 (2.4%) |  |
|  | Not reported | 13 (5.8%) | 491 (8.0%) |  |
| RV Function | Normal | 187 (83.1%) | 4289 (78.7%) | <0.001 |
|  | Borderline | 3 (1.3%) | 4 (0.1%) |  |
|  | Mildly reduced | 17 (7.6%) | 436 (7.1%) |  |
|  | Mild to moderately reduced | 3 (1.3%) | 134 (2.2%) |  |
|  | Moderately reduced | 3 (1.3%) | 0 (0.0%) |  |
|  | Mod. to severely reduced | 1 (0.4%) | 0 (0.0%) |  |
|  | Severely reduced | 0 (0.0%) | 7 (0.1%) |  |
|  | Not reported | 11 (4.9%) | 730 (11.9%) |  |
| TAPSE (cm), mean±SD |  | 2.3±1.1<br>(n=172) | 2.5±1.5<br>(n=5,887) | 0.22 |
| TAPSE <1.7, n (%) |  | 26 (15.1%) | 1,205 (20.5%) | 0.09 |
| RV S' velocity, cm/s, mean±SD |  | 14.4±4.2<br>(n=78) | 13.6±4.4<br>(n=2,936) | 0.11 |
| RV S' <9.5cm/s |  | 8 (10.3%) | 468 (15.9%) | 0.17 |
| TR peak gradient, mean±SD |  | 31.1±12.8<br>(n=73) | 34.2±14.9<br>(n=3,386) | 0.074 |
| TR peak gradient ≥ 2.9m/s |  | 26 (35.6%) | 1623 (47.9%) | 0.044 |
| RV Systolic Pressure (mm Hg), mean±SD |  | 35.7±14.6 | 39.6±16.5 | 0.029 |
| Right Atrial Pressure (mmHg), mean±SD |  | 4.6±3.5<br>(n=103) | 5.6±7.9<br>(n=4,338) | 0.18 |
| RV s'/RVSP ratio |  | 0.44±0.31<br>(n=40) | 0.39±0.20<br>(n=1,730) | 0.16 |
| TAPSE/RVSP ratio |  | 0.071±0.03<br>(n=84) | 0.070±0.05<br>(n=3,331) | 0.97 |

### RV Dilation and Reduced RV Function are Associated with Higher In-Hospital Mortality in COVID and Non-COVID Respiratory Illness

Among those with COVID-19, 26 (16.5%) with normal RV size, 8 (27.6%) with mild RV dilation, and 5 (41.7%) with moderate to severe RV dilation died. Similarly, among those with other respiratory illness, 441 (10.0%) with normal RV size, 115 (14.3%) with mild RV dilation, and 88 (19.7%) of those with moderate to severe RV dilation died. Among those with COVID-19, 36 (19%) with normal RV function, 6 (30%) with mild RV dysfunction, and 2 (29%) with greater than mild RV dysfunction died. Among the control group, 487 (10%) with normal RV function, 67 (15.2%) with mild RV dysfunction, and 25 (18%) with more than mild RV dysfunction died.

In adjusted analyses, independent of COVID-19, mild RV dilation was associated with 1.41 times higher relative risk of inpatient mortality (95%CI 1.17 to 1.69; p=0.0003) and moderate to severe RV dilation with 2.00 times higher risk of inpatient mortality (95%CI 1.62 to 2.47; p<0.0001), or absolute risk differences of 4.2% and 10.3% higher mortality, respectively. Similarly, mild RV dysfunction was associated with 1.39 times higher relative risk of inpatient mortality (95%CI 1.10 to 1.77; p=0.007) and greater than mild dysfunction with 1.68 times higher risk of inpatient mortality (95%CI 1.17 to 2.42; p=0.005), or absolute risk differences of 4.1% and 7.1% higher mortality, respectively. Results were similar when those with history of pulmonary hypertension were excluded (not shown).

As shown in Supplemental Table 1, the relative risk of mortality associated with RV dilation and RV dysfunction among those with COVID-19 was similar to the relative risk among those with other respiratory illness. Because the baseline risk is higher among those with COVID-19, the similar relative risks translate into greater absolute risk increase associated with RV dilation and RV dysfunction among those with COVID-19 and RV dilation or dysfunction compared to those without COVID-19 (Table 3).

**Table 3.** Relative Risk by COVID and RV Status Compared to Other Respiratory Illness with Normal RV Adjusted relative risk by COVID-19 and RV Size/Function adjusted for age, sex, past medical history, admission hemoglobin and eGFR, and intubation at the time of echocardiogram. The confidence intervals for RV dysfunction amogn the COVID+ were wide due to the small numbers but fit with the same pattern of a dose response as other respiratory illness.

|  | **Non-COVID-19 Acute**<br>**Respiratory Illness** | **COVID-19** |
| --- | --- | --- |
| **Normal RV size** | 1.00 (REF) | 1.55 (95% CI 1.08, 2.22) |
| **RV mildly dilated** | 1.42 (95% CI 1.17, 1.71) | 1.90 (95% CI 0.98, 3.67) |
| **RV ≥ moderately dilated** | 2.03 (95% CI 1.64, 2.50) | 3.00 (95% CI 1.39, 6.46) |
| **Normal RV Function** | 1.00 (REF) | 1.50 (95% CI 1.08, 2.07) |
| **RV mild dysfunction** | 1.41 (95% CI 1.11, 1.80) | 1.65 (95% CI 0.66, 4.10) |
| **RV > mild dysfunction** | 1.67 (95% CI 1.16, 2.40) | 2.66 (95% CI 0.87, 8.15) |

Although RV size and function are correlated (r=0.42, p<0.0001), there were no significant multiplicative interactions between RV function and RV volume on in-hospital mortality (p_interaction_=0.96) or COVID-19 status and RV dilation or RV dysfunction on mortality (p_interaction_=0.92 and 0.97, respectively). Adding RV function to the model that included RV volume was statistically significantly better (p=0.02), but adding RV volume to the model that included RV function was not statistically significantly better (p=0.11). Accounting for RV dilation and function together, those with RV dysfunction alone have 36% higher risk (95%CI 0.99 to 1.88; p=0.06), RV dilation alone have an 66% higher risk (95%CI 1.40 to 1.97; p<0.0001), and those with both RV dilation and dysfunction have 68% higher risk (1.30 to 2.17; p<0.0001) relative to normal RV size and function.

As shown in Figure 1a, by Fine and Gray models adjusted for the same covariates, among those remaining hospitalized at a given length of stay, mild RV dilation was associated with 47% higher hazard of mortality (SHR 1.47, 95%CI 1.19-1.80; p<0.001) and moderate to severe RV dilation was associated with 120% higher mortality hazard (SHR 2.20; 95%CI 1.71-2.84; p<0.001) compared to normal RV size. Accounting for RV size, COVID-19 was associated with 51% higher hazard of mortality (SHR 1.51, 95%CI 1.09 to 2.12; p=0.01) compared to those without COVID-19. Similarly, mild RV dysfunction was associated with 47% higher mortality hazard (SHR 1.47; 95%CO 1.12 to 1.93; p=0.005) and moderate or greater RV dysfunction was associated with 83% higher hazard of mortality (SHR 1.19; 95%CI 1.19 to 2.81; p=0.006). Accounting for RV function, COVID-19 was associated with 47% higher risk of mortality (SHR 1.47; 95%CI 1.04 to 2.07; p=0.03).

**Figure 1.**
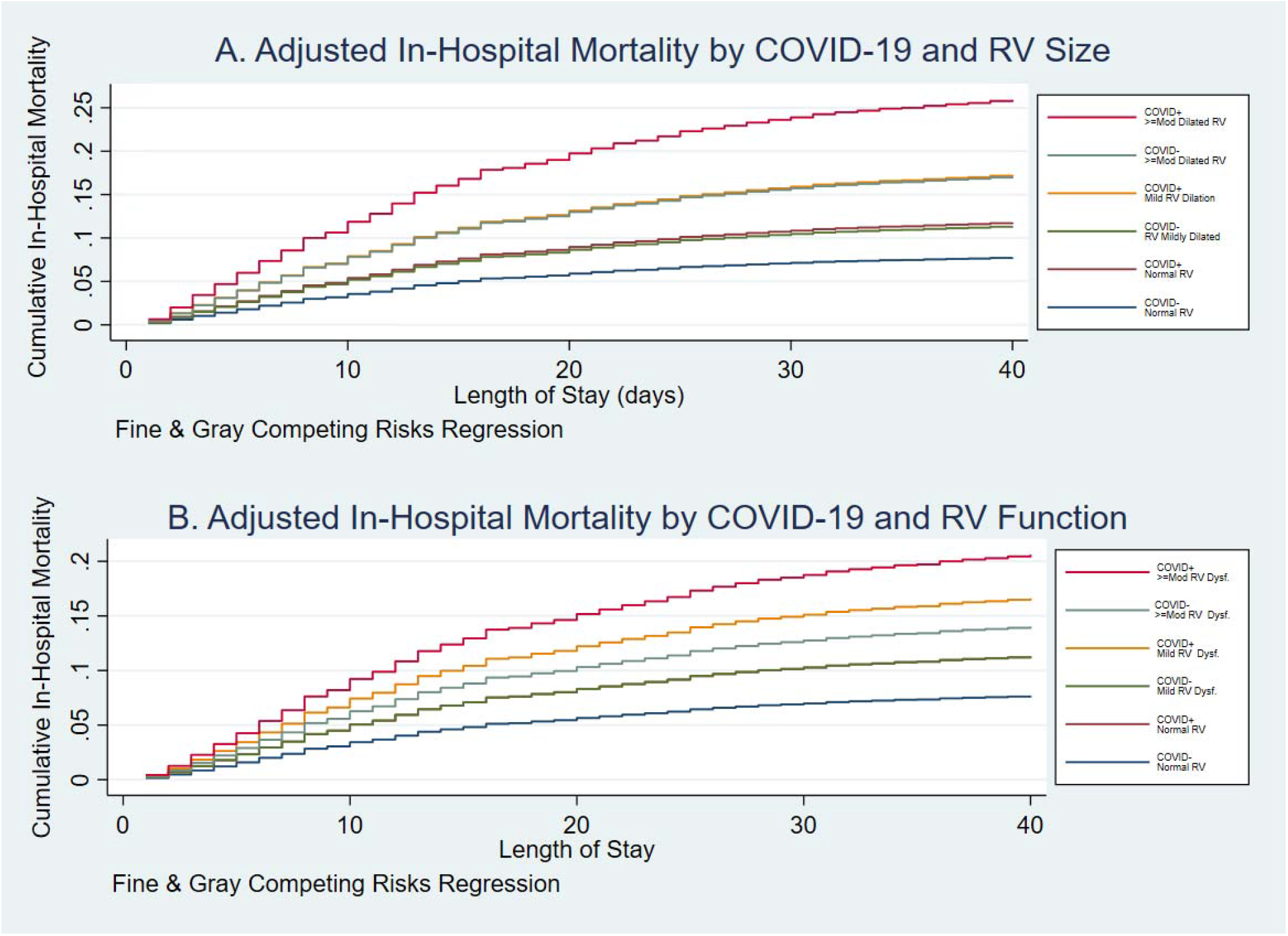
Adjusted In Hospital Mortality by COVID-19 and RV Size and Function Figure 1: Cumulative incidence functions of adjusted in-hospital mortality by COVID-19 status and RV size (A, top) and RV function (B, bottom) using Fine & Gray competing risk regression to account for discharge home. Model is adjusted for age, sex, race/ethnicity, past medical history, admission hemoglobin and creatinine, and intubation at the time of the echocardiogram.

### Cardiac Biomarkers, RV Dilation, and Tricuspid Regurgitation Gradient as Predictors of Mortality

Among those with respiratory illness, the ordinal variable for RV size has an AUROC of 0.56 for in-hospital mortality and RV function has an AUROC of 0.53. Table 4 demonstrates that with incremental worsening of RV dilation and dysfunction, there is a similar increase in specificity and positive likelihood ratio for in-hospital mortality.

**Table 4.** Sensitivity, Specificity, Classification Accuracy, and Likelihood Ratios for In Hospital Mortality Sensitivity, specificity, accuracy, positive and negative liklihood ratios for inhospital mortality by qualitative RV volume and function assessment.

| Cutpoint | Sensitivity | Specificity | Mortality<br>Correctly<br>Classified | LR+ | LR- |
| --- | --- | --- | --- | --- | --- |
| *RV Volume* |  |  |  |  |  |
| ≥Normal | 100% | 0% | 11.8% | 1.000 |  |
| ≥Borderline Dilated | 31.7% | 79.2% | 73.6% | 1.53 | 0.86 |
| ≥Mildly Dilated | 28.0% | 82.4% | 76.0% | 1.59 | 0.87 |
| ≥Mild-Moderately Dilated | 17.0% | 90.7% | 82.0% | 1.82 | 0.92 |
| ≥Moderately Dilated | 13.5% | 92.9% | 83.6% | 1.90 | 0.93 |
| ≥Moderately to Severely Dilated | 6.1% | 96.8% | 86.2% | 1.91 | 0.97 |
| ≥Severely Dilated | 4.6% | 97.7% | 86.7% | 1.99 | 0.97 |
| >Severely Dilated | 0% | 100% | 88.3% |  | 1.00 |
| *RV Function* |  |  |  |  |  |
| ≥Normal | 100% | 0% | 11.1% | 1.000 |  |
| ≥Borderline Dysfunction | 16.1% | 89.9% | 81.7% | 1.58 | 0.93 |
| ≥Mild Dysfunction | 15.4% | 89.9% | 81.7% | 1.53 | 0.94 |
| ≥Mild-Moderate Dysfunction | 4.33% | 97.6% | 87.3% | 1.79 | 0.98 |
| ≥Moderate Dysfunction | 0.8% | 99.9% | 88.9% | 6.70 | 0.99 |
| ≥Moderate to Severe Dysfunction | 0.64 | 99.9% | 88.9% | 8.04 | 0.99 |
| ≥Severe Dysfunction | 0.64% | 99.9% | 88.9% | 10.72 | 0.99 |
| >Severe Dysfunction | 0% | 100% | 88.9% |  | 1.00 |

Compared to a basic model that included age, sex, race/ethnicity, body mass index, past medical history, admission hemoglobin and eGFR, COVID diagnosis, and mechanical ventilation at the time of the echocardiogram (AUROC=0.71), adding cardiac biomarkers (0.72; p=0.005), RV dilation (0.72, p=0.02), RV dysfunction (0.73, p=0.05), or RV size, function and biomarkers (0.74, p=0.002) slightly improved the model (Figure 2a). Compared to the model with RV size, function, and biomarkers, adding TR gradient significantly increased the AUROC (0.70 to 0.74; p=0.002), but incorporating LVEF (0.74 to 0.74, p=0.79) or TAPSE (0.74 to 0.75; p=0.12) did not. Among 845 with RV s’ reported (which may select for those at higher risk), including RV s’ dramatically improved the AUROC compared to the model with qualitative RV size and function and cardiac biomarkers (0.71 to 0.79; p=0.001, Figure 2b) and even compared to the model incorporating qualitative RV size and function, cardiac biomarkers, LVEF, TR gradient, and TAPSE (0.75 to 0.79; p=0.009). Incorporation of RV/PA coupling assessed by TAPSE/RVSP ratio or RV s’/RVSP ratio did not improve the AUROC (p=0.06 and p=0.39) comparing to including TAPSE, RV s’, and tricuspid gradient independently. Adding both qualitative and quantitative RV size and function variables resulted in improvement in the AUROC from 0.69 to 0.79 (p=0.0008), an integrated discrimination improvement of 9.8% (p<0.00001) and a net reclassification index of 40% using cutpoints of 5%, 10%, and 15% probability of mortality (roughly quartiles for total sample) or 34% with cutpoints of 10%, 20%, and 30% (roughly quartiles for the COVID-19 population; p<0.0001).

**Figure 2.**
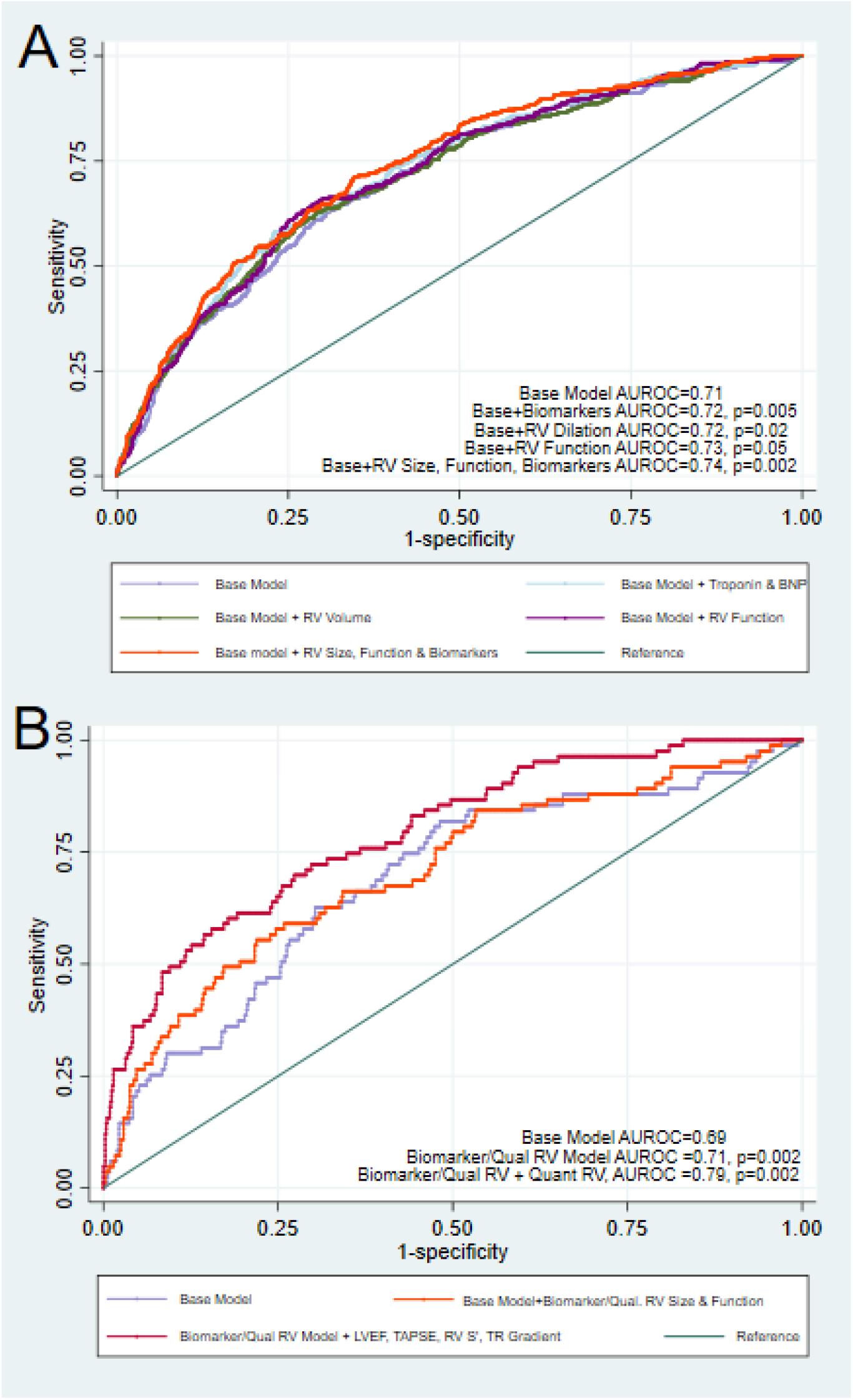
Receiver-Operator Curves for Prediction Models for In Hospital Mortality Figure 2: In panel A, we present the receiver operative curves for models to predict inpatient mortality with the addition of biomarkers and qualitative RV size and function or both, all of which resulted in modest but statistically significant improvements in the AUROC compared to the base model (p-values shown are all compared to the base model). As shown in panel B, incorporating quantitative parameters in addition to qualitative parameters increased the AUROC among those with complete data (n=845).

### Quantitative RV Assessment of RV Size vs Qualitative Assessments

In terms of quantitative RV measurements, there was an 81.2% agreement between readers for the measurement of the RV basal diameter (ICC=0.84, CV=12.6; kappa=0.54 for the binary “dilated”), and an 82.3% agreement for FAC (ICC=0.80, CV=19; kappa=0.57 for RV dysfunction by FAC). Overall, there was highly statistically significant (p<0.001) associations between the qualitative and quantitative measurements of RV size and function with significant overlap between the two (Figure 3, Supplemental table 2).

**Figure 3.**
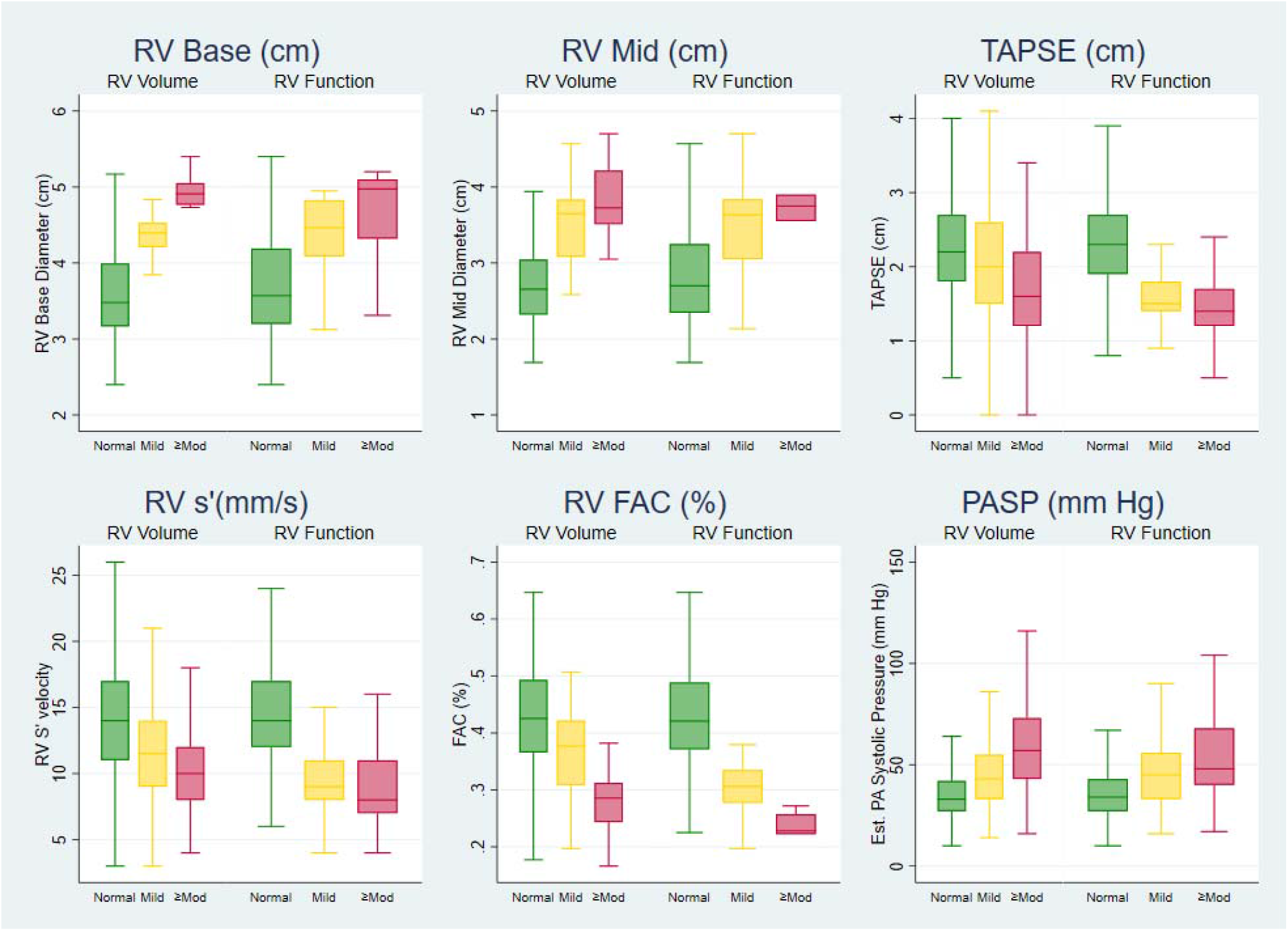
Comparison of RV Quantitative and Qualitative Parameters Among Individuals with COVID-19 Figure 3 Legend: Box and whisker plots of RV quantitative parameters including RV base diameter, RV mid diameter, TAPSE, RV s’, RV FAC, and PASP by qualitative RV dilation and RV dysfunction categories.

In unadjusted analyses among those with COVID-19, larger RV mid-cavity diameter was associated with in-hospital mortality (p=0.03), but not RV basal diameter (p=0.93), RV end diastolic area (p=0.65), RV end systolic area (p=0.89), or Fractional Area Change (p=0.42). After adjustment, RV dimensions were not associated with in-hospital mortality. Among those with COVID-19, incorporating RV dimensions into the predictive models did not significantly improve the AUROC (p=0.78 for basal diameter, p=0.79 for mid diameter, p=0.14 for end diastolic area, p=0.31 for end systolic area, p=0.79 for FAC). Inability to perform RV measurements due to poor RV free wall visualization, however, was associated with mortality (ARR 1.92; 95%CI 1.19 to 3.11; p=0.008).

### RV Dilation is Associated with Pulmonary Embolism, Subsequent Mechanical Ventilation, and Incident Atrial Fibrillation

Among those without prior history of venous thromboembolism, mild RV dilation was associated with 1.36 times higher relative risk of DVT or PE diagnosed during the hospitalization (95%CI 1.11 to 1.65) and 1.69 times higher risk of PE (95%CI 1.28 to 2.23; p=0.0002). Moderate to severe RV dilation was associated with 1.46 times higher relative risk of diagnosed DVT or PE (95%CI 1.15 to 1.86; p=0.002) and 2.10 times higher risk of PE (95%CI 1.53 to 2.88; p<0.0001). Mild RV dysfunction was not associated with VTE or PE, but moderate-to-severe dysfunction was associated with VTE (ARR 1.62; 95%CI 1.14 to 2.29; p=0.007) and PE (ARR 2.39; 95%CI 1.55 to 3.70; p=0.0001).

Among patients who were not intubated at the time of their echocardiogram, RV dilation was associated with 1.15 times higher risk of subsequently requiring intubation and mechanical ventilation (95%CI 1.00 to 1.32; p=0.05), but RV dysfunction was not (p=0.17). Results were similar among those with COVID-19 compared to those with other respiratory illness.

RV dilation was also associated with development of incident atrial fibrillation among those without a history of atrial fibrillation or atrial fibrillation present on their admitting electrocardiogram or echocardiogram, with an adjusted relative risk of 1.46 (95%CI 1.24 to 1.73; p<0.0001) for mild RV dilation and 1.72 (95%CI 1.40 to 2.10; p<0.0001) for moderate or greater RV dilation, with no difference by COVID-19 status (p_interaction_=0.74). Similarly, RV dysfunction was also associated with incident atrial fibrillation (ARR 1.61; 95%CI 1.34 to 1.92).

Moderate to severe RV dilation was highly associated with initiation of renal replacement therapy among those with COVID-19 not previously on hemodialysis (ARR 3.85, 95%CI 1.54-9.66; p=0.004), but not to a statistically significant degree among those with other respiratory illness (ARR 1.32, 95%CI 0.82-2.12; p=0.25). Mild RV dilation was not associated with risk of initiation of renal replacement therapy, and RV dysfunction was not statistically significant (p=0.06).

## DISCUSSION

In our study of 225 adults hospitalized with acute COVID-19 and 6,150 adults hospitalized with influenza, pneumonia or ARDS who underwent clinical inpatient echocardiography, the presence of RV dilation or RV dysfunction was associated with higher risk of in-hospital mortality in both acute COVID-19 and other respiratory illness. Those with COVID-19 had a higher risk of inpatient mortality than the control population admitted for non-COVID respiratory illness. The relative risk associated with RV dilation or RV dysfunction was similar among those with COVID-19 compared to other acute respiratory illness. Incorporation of RV s’ but not other RV quantitative measurements significantly improved predictive model performance compared to qualitative assessment. RV dilation is associated with diagnosis of venous thromboembolism, subsequent intubation and mechanical ventilation, and diagnosis of atrial fibrillation, although these should be interpreted with caution given the possibility of timing misclassification from EHR records.

To our knowledge, this study is one of the first to compare RV dilation and dysfunction and their associations with mortality in those with acute COVID-19 to a population of patients admitted for other respiratory illness. The prevalence of RV dilation (21%) in our cohort is similar to that reported by a recent meta-analysis of RV abnormalities in acute COVID-19 (15-40%).^13^ However, RV dysfunction was less prevalent in our study (7%) than in the meta-analysis (13-46%).^13^ Our findings add to the growing evidence that RV dilation and dysfunction portend worse outcomes in those admitted for acute COVID-19, especially in those exhibiting both dilation and dysfunction on echocardiogram.^12,13^

In prior studies of adults admitted for H1N1 influenza, RV dilation and dysfunction were common echo findings, but neither was found to be associated with mortality.^19,20^ There is mixed evidence surrounding the prognostic implications of RV systolic dysfunction in ARDS from non-COVID etiologies, and even less is known about the significance of RV dilation without dysfunction.^21,22,23,24^ We conclude from our study that both RV dilation and dysfunction are associated with in-hospital mortality in those admitted for influenza, pneumonia or ARDS, which highlights the key role played by the RV in acute pulmonary disease.

The pathophysiology of RV dysfunction in ARDS stem from numerous insults to the pulmonary circulation, including mechanical compression by interstitial edema, hypoxic-induced pulmonary vasoconstriction and microvascular thrombosis.^25^ In addition to causing RV dilation and dysfunction, these conditions as well as hypercoagulability from acute illness predispose patients with acute respiratory illness to macrovascular obstruction in the form of acute PE, which can also lead to subsequent RV remodeling.^26^ In addition, high PEEP in mechanical ventilation further increases the compression of alveolar vessels and subsequently raises the pulmonary vascular resistance (PVR). As the RV is vulnerable to even slight increases in PVR, the increase in RV afterload leads to an uncoupling of RV systolic function and the pulmonary circulation, leading to RV dilation and dysfunction.^27^ Given the importance of the RV in acute pulmonary disease, there has been a recent push for ventilation strategies to prevent acute cor pulmonale and reduce mortality by keeping plateau pressure less than 17mmHg.^28^ Our finding of the association between RV dilation and mortality in those admitted for influenza, bacterial pneumonia and ARDS warrants further investigation of the prognostic utility of finding early RV dilation in hypoxic lung disease.

RV size and function can be challenging to assess by echocardiogram, especially in technically difficult inpatient portable studies. Interestingly, among those with RV s’ measurements, including RV s’ significantly improved the AUROC in contrast to TAPSE. Quantitative measurements of RV size did not outperform qualitative assessment among those with COVID-19. This is possibly because, unlike the LV, the RV has a complex asymmetric crescent shape and its retro-sternal position often precludes accurate measurement.^29,30^ As a result, current guidelines recommend combining a qualitative and quantitative approach to assessing RV function by reporting one or more of FAC, TAPSE or RV S’.^17^ Furthermore, assessment of the RV is especially difficult in intubated ICU patients, where obtaining adequate RV-focused views is even more challenging. In fact, we found that those with poorly visualized RV free wall were at higher risk of mortality compared to those with adequate views on echo. A significant portion of the critically-ill population with COVID-19 was excluded from the quantitative portion of our study despite being at the highest risk of death, which may contribute to the lack of association found between quantitative RV dilation and mortality. Although we demonstrated that visual measurements of RV size and function correlated well with objective measurements, there is still a need for further studies to examine the level of congruence between qualitative and quantitative assessment of the RV, where there is currently mixed evidence.^31,32^

### Clinical Implications and Future Directions

The prognostic significance of RV dilation and dysfunction among those admitted for respiratory illness raises questions as to whether a “RV protective” ventilation strategy may reduce mortality. In addition, as RV dilation was associated with the need for mechanical ventilation, the presence of dilation on echocardiogram may serve as an early prognostic tool in predicting the course of respiratory illness. The fact that those hospitalized for acute COVID-19 had a higher mortality rate than our control population despite having a significantly lower PASP suggests that there are mechanisms by which COVID-19 causes mortality beyond increased PVR.

An avenue for future assessment of the RV is the incorporation of RV longitudinal strain (RVLS) and 3D echocardiography, which has the ability to provide more sensitive and accurate measurements of RV size and function compared to conventional 2D echocardiography alone.^33,34^ There is evidence that both reduced RVLS and RVEF on 3D echocardiography were associated with worse outcomes in COVID-19^9,35^, but it is unclear if these modalities provide prognostic value in respiratory disease from non-COVID etiologies. Although we did not compare pre-illness echocardiograms (which would provide stronger evidence of causality), others have demonstrated that RV dilation and dysfunction after acute COVID-19 compared to prior echocardiograms without those finds is associated with mortality.^36^

### Limitations

The primary limitation is that this study was conducted primarily through retrospective review of electronic health records, which carries risk of misclassification. However, through manual chart review, we clinically adjudicated all COVID-19 cases and did not find any misclassification of the primary outcome of in-hospital mortality and manually excluded the few hospitalizations during this period of patients with positive SARS-CoV-2 PCR without clinical evidence of COVID-19. Follow-up was limited to the time of discharge which is why we used a competing risk regression to account for discharge alive. We extracted the echocardiographic reports for echocardiograms performed during the hospitalization with additional measurements performed on all echocardiograms of COVID-19 but not control participants. Qualitative assessment of the RV were performed clinical echocardiographers who were not blinded to the clinical status of the patient, although in practice few readers open the clinical charts. Since we did not compare to prior echocardiograms, we could not exclude those with pre-existing RV dilation or dysfunction; we did exclude those with a history of pulmonary arterial hypertension in a sensitivity analysis with similar results. The control group represents a heterogenous population admitted for a variety of acute pulmonary disease, but we chose this control group to be similar to those with COVID-19 in terms of unmeasured confounders. External validity may be limited as the UCSF Health hospitals reside in a single metropolitan area and echocardiograms were interpreted by UCSF cardiologists. Direct measurements of the RV were at times limited by poor windows and strain imaging was not feasible in many participants. To be included, the patients had to have a clinically indicated echocardiogram as judged by the inpatient team, so selection bias limits estimation of the prevalence of these echocardiographic findings to those who had a clinical echocardiogram performed. We did not use machine learning approaches for optimizing prediction models, nor did we test them through bootstrapping or external validation cohorts as our goal was to demonstrate prognostic utility rather than develop and validate prediction models.

### Conclusions

In those hospitalized for COVID-19 and in those with influenza, pneumonia or ARDS, echocardiographic evidence of RV dilation and RV dysfunction were associated with in-hospital mortality, with the worst prognosis in those with both COVID-19 and RV dilation or dysfunction. RV dilation is associated with diagnosis of venous thromboembolism, diagnosis of atrial fibrillation, and subsequent intubation and mechanical ventilation.

## Data Availability

Deidentified data may be available upon reasonable request to the authors and legal approval from UCSF.

## ACKNOWLEDGEMENTS

The authors wanted to acknowledge the assistance provided by the UCSF CTSI in the collection of clinical patient data, including laboratory and imaging results. This publication was supported by the National Center for Advancing Translational Sciences, National Institutes of Health, through UCSF-CTSI Grant Number UL1 TR001872. Its contents are solely the responsibility of the authors and do not necessarily represent the official views of the NIH.

---

**Supplemental Table 1.** Adjusted Relative Risk for RV Size & Function Conditional on COVID Status The estimated adjusted relative risk by RV size and function categories was similar among those with COVID-19 and among the controls with other respiratory illness consistent with the non-significant p-values for interaction.

|  | Overall ARR | ARR COVID+ | ARR COVID - | P interaction |
| --- | --- | --- | --- | --- |
| Normal RV size | 1.00 (ref) | 1.00 (ref) | 1.00 (ref) | 0.92 |
| RV mildly dilated | 1.41 (95%CI 1.17 to 1.69; p=0.0003) | 1.37 (95% CI 1.16, 1.62; p=0.0003) | 1.41 (95% CI 1.17, 1.69; p<0.0001) |  |
| RV ≥moderately dilated | 2.00 (95%CI 1.62 to 2.47; p<0.0001) | 1.88 (95%CI 1.55, 2.28; p<0.0001) | 2.01 (95% CI 1.63 2.48; p<0.0001) |  |
| Normal RV Function | 1.00 (ref) | 1.00 (ref) | 1.00 (ref) | 0.97 |
| RV mild dysfunction | 1.39 (95%CI 1.10 to 1.77; p=0.007) | 1.36 (95%CI 1.09, 1.69; p=0.006) | 1.39 (95% CI 1.10, 1.77; p=0.007) |  |
| RV > mild dysfunction | 1.68 (95%CI 1.17 to 2.42; p=0.005) | 1.61 (95% CI 1.16, 2.26; p=0.005) | 1.69 (95% CI 1.17, 2.44; p=0.006) |  |

**Supplemental Table 2:**
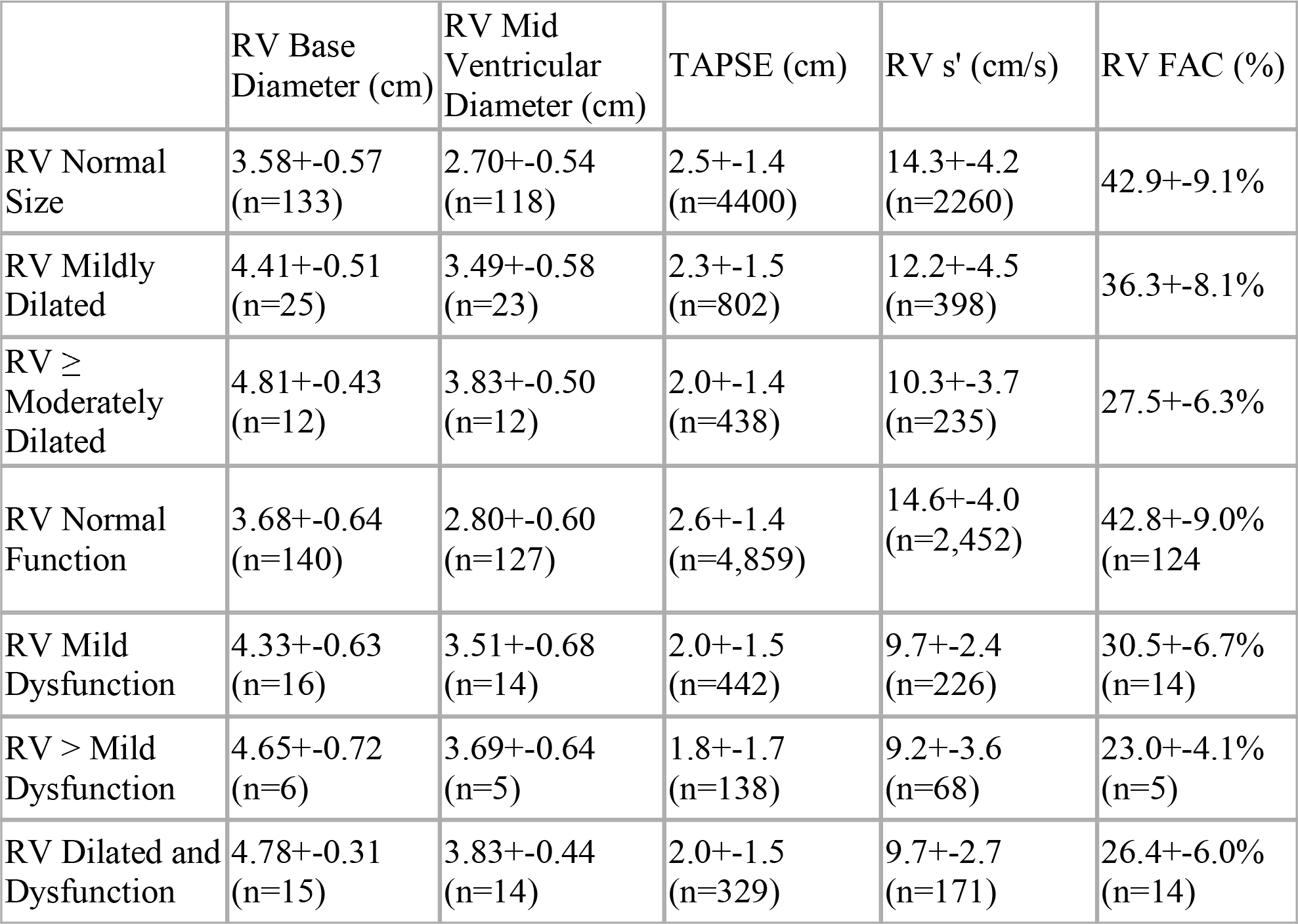
Correlation Between Qualitative (Rows) and Quantitative (Columns) Measurements of RV Size and Function Qualitative measurements of RV size and function correlates well with quantitative measurements. All exhibited a highly statistically significant association (p < 0.001) across the RV size categories and across the RV function categories, except for RV mid for RV function >mildly dysfunction (p=0.13) and RV base for RV function >mild (p<0.03). Poor RV view include those where quantitative measurements of the RV could not be accurately made, which were ∼30% for all categories.

|  | RV Base Diameter (cm) | RV Mid Ventricular Diameter (cm) | TAPSE (cm) | RV s' (cm/s) | RV FAC (%) |
| --- | --- | --- | --- | --- | --- |
| RV Normal Size | 3.58+-0.57 (n=133) | 2.70+-0.54 (n=118) | 2.5+-1.4 (n=4400) | 14.3+-4.2 (n=2260) | 42.9+-9.1% |
| RV Mildly Dilated | 4.41+-0.51 (n=25) | 3.49+-0.58 (n=23) | 2.3+-1.5 (n=802) | 12.2+-4.5 (n=398) | 36.3+-8.1% |
| RV ≥ Moderately Dilated | 4.81+-0.43 (n=12) | 3.83+-0.50 (n=12) | 2.0+-1.4 (n=438) | 10.3+-3.7 (n=235) | 27.5+-6.3% |
| RV Normal Function | 3.68+-0.64 (n=140) | 2.80+-0.60 (n=127) | 2.6+-1.4 (n=4,859) | 14.6+-4.0 (n=2,452) | 42.8+-9.0% (n=124) |
| RV Mild Dysfunction | 4.33+-0.63 (n=16) | 3.51+-0.68 (n=14) | 2.0+-1.5 (n=442) | 9.7+-2.4 (n=226) | 30.5+-6.7% (n=14) |
| RV > Mild Dysfunction | 4.65+-0.72 (n=6) | 3.69+-0.64 (n=5) | 1.8+-1.7 (n=138) | 9.2+-3.6 (n=68) | 23.0+-4.1% (n=5) |
| RV Dilated and Dysfunction | 4.78+-0.31 (n=15) | 3.83+-0.44 (n=14) | 2.0+-1.5 (n=329) | 9.7+-2.7 (n=171) | 26.4+-6.0% (n=14) |

## REFERENCES

1. Lala A, Johnson KW, Januzzi JL, et al. Prevalence and Impact of Myocardial Injury in Patients Hospitalized With COVID-19 Infection. J Am Coll Cardiol. 2020;76(5):533–546. doi:10.1016/j.jacc.2020.06.007

2. Shi S, Qin M, Cai Y, et al. Characteristics and clinical significance of myocardial injury in patients with severe coronavirus disease 2019. Eur Heart J. 2020;41(22):2070–2079. doi:10.1093/eurheartj/ehaa408

3. Shi S, Qin M, Shen B, et al. Association of Cardiac Injury With Mortality in Hospitalized Patients With COVID-19 in Wuhan, China. JAMA Cardiol. 2020;5(7):802–810. doi:10.1001/jamacardio.2020.0950

4. Szekely Y, Lichter Y, Taieb P, et al. Spectrum of Cardiac Manifestations in COVID-19: A Systematic Echocardiographic Study. Circulation. 2020;142(4):342–353. doi:10.1161/CIRCULATIONAHA.120.047971

5. Mahmoud-Elsayed HM, Moody WE, Bradlow WM, et al. Echocardiographic Findings in Patients With COVID-19 Pneumonia. Can J Cardiol. 2020;36(8):1203–1207. doi:10.1016/j.cjca.2020.05.030

6. Repessé X, Charron C, Vieillard-Baron A. Right ventricular failure in acute lung injury and acute respiratory distress syndrome. Minerva Anestesiol. 2012;78(8):941–948.

7. Osman D, Monnet X, Castelain V, et al. Incidence and prognostic value of right ventricular failure in acute respiratory distress syndrome. Intensive Care Med. 2009;35(1):69–76. doi:10.1007/s00134-008-1307-1

8. Sato, R., Dugar, S., Cheungpasitporn, W. et al. The impact of right ventricular injury on the mortality in patients with acute respiratory distress syndrome: a systematic review and meta-analysis. Crit Care 25, 172 (2021). https://doi.org/10.1186/s13054-021-03591-9

9. Li Y, Li H, Zhu S, et al. Prognostic Value of Right Ventricular Longitudinal Strain in Patients With COVID-19. JACC Cardiovasc Imaging. 2020;13(11):2287–2299. doi:10.1016/j.jcmg.2020.04.014

10. Karagodin I, Carvalho Singulane C, Woodward GM, et al. Echocardiographic Correlates of In-Hospital Death in Patients with Acute COVID-19 Infection: The World Alliance Societies of Echocardiography (WASE-COVID) Study. J Am Soc Echocardiogr. 2021;34(8):819–830. doi:10.1016/j.echo.2021.05.010

11. Zuin M, Rigatelli G, Roncon L, Zuliani G. Relationship between echocardiographic tricuspid annular plane systolic excursion and mortality in COVID-19 patients: A Meta-analysis. Echocardiography. 2021;38(9):1579–1585. doi:10.1111/echo.15175

12. Argulian E, Sud K, Vogel B, et al. Right Ventricular Dilation in Hospitalized Patients With COVID-19 Infection. JACC Cardiovasc Imaging. 2020;13(11):2459–2461. doi:10.1016/j.jcmg.2020.05.010

13. Paternoster G, Bertini P, Innelli P, et al. Right Ventricular Dysfunction in Patients With COVID-19: A Systematic Review and Meta-analysis. J Cardiothorac Vasc Anesth. 2021;35(11):3319–3324. doi:10.1053/j.jvca.2021.04.008

14. Chotalia M, Ali M, Alderman JE, et al. Right Ventricular Dysfunction and Its Association With Mortality in Coronavirus Disease 2019 Acute Respiratory Distress Syndrome. Crit Care Med. 2021;49(10):1757–1768. doi:10.1097/CCM.0000000000005167

15. Schott JP, Mertens AN, Bloomingdale R, et al. Transthoracic echocardiographic findings in patients admitted with SARS-CoV-2 infection. Echocardiography. 2020;37(10):1551–1556. doi:10.1111/echo.14835

16. Rath D, Petersen-Uribe Á, Avdiu A, et al. Impaired cardiac function is associated with mortality in patients with acute COVID-19 infection. Clin Res Cardiol. 2020;109(12):1491–1499. doi:10.1007/s00392-020-01683-0

17. Rudski LG, Lai WW, Afilalo J, et al. Guidelines for the echocardiographic assessment of the right heart in adults: a report from the American Society of Echocardiography endorsed by the European Association of Echocardiography, a registered branch of the European Society of Cardiology, and the Canadian Society of Echocardiography. J Am Soc Echocardiogr. 2010;23(7):685–788. doi:10.1016/j.echo.2010.05.010

18. Oulhaj A, Ahmed LA, Prattes J, et al. The competing risk between in-hospital mortality and recovery: A pitfall in COVID-19 survival analysis research. medRxiv 2020.07.11.20151472; doi: https://doi.org/10.1101/2020.07.11.20151472

19. Brown SM, Pittman J, Miller Iii RR, et al. Right and left heart failure in severe H1N1 influenza A infection. Eur Respir J. 2011;37(1):112–118. doi:10.1183/09031936.00008210

20. Fagnoul D, Pasquier P, Bodson L, Ortiz JA, Vincent JL, De Backer D. Myocardial dysfunction during H1N1 influenza infection. J Crit Care. 2013;28(4):321–327. doi:10.1016/j.jcrc.2013.01.010

21. Wadia SK, Shah TG, Hedstrom G, Kovach JA, Tandon R. Early detection of right ventricular dysfunction using transthoracic echocardiography in ARDS: a more objective approach. Echocardiography. 2016;33(12):1874–1879. doi:10.1111/echo.13350

22. Osman D, Monnet X, Castelain V, et al. Incidence and prognostic value of right ventricular failure in acute respiratory distress syndrome. Intensive Care Med. 2009;35(1):69–76. doi:10.1007/s00134-008-1307-1

23. Lhéritier G, Legras A, Caille A, et al. Prevalence and prognostic value of acute cor pulmonale and patent foramen ovale in ventilated patients with early acute respiratory distress syndrome: a multicenter study. Intensive Care Med. 2013;39(10):1734–1742. doi:10.1007/s00134-013-3017-6

24. Vieillard-Baron A, Schmitt JM, Augarde R, et al. Acute cor pulmonale in acute respiratory distress syndrome submitted to protective ventilation: incidence, clinical implications, and prognosis [published correction appears in Crit Care Med 2002 Mar;30(3):726]. Crit Care Med. 2001;29(8):1551–1555. doi:10.1097/00003246-200108000-00009

25. Zochios V, Parhar K, Tunnicliffe W, Roscoe A, Gao F. The Right Ventricle in ARDS. Chest. 2017;152(1):181–193. doi:10.1016/j.chest.2017.02.019

26. Pinsky MR. The right ventricle: interaction with the pulmonary circulation [published correction appears in Crit Care. 2016 Nov 10;20(1):364]. Crit Care. 2016;20(1):266. Published 2016 Sep 10. doi:10.1186/s13054-016-1440-0

27. Spruijt OA, de Man FS, Groepenhoff H, et al. The effects of exercise on right ventricular contractility and right ventricular-arterial coupling in pulmonary hypertension. Am J Respir Crit Care Med. 2015;191(9):1050–1057. doi:10.1164/rccm.201412-2271OC

28. Jardin F, Vieillard-Baron A. Is there a safe plateau pressure in ARDS? The right heart only knows. Intensive Care Med. 2007;33(3):444–447. doi:10.1007/s00134-007-0552-z

29. Karas MG, Kizer JR. Echocardiographic assessment of the right ventricle and associated hemodynamics. Prog Cardiovasc Dis. 2012;55(2):144–160. doi:10.1016/j.pcad.2012.07.011

30. Dutta T, Aronow WS. Echocardiographic evaluation of the right ventricle: Clinical implications. Clin Cardiol. 2017;40(8):542–548. doi:10.1002/clc.22694

31. Ling LF, Obuchowski NA, Rodriguez L, Popovic Z, Kwon D, Marwick TH. Accuracy and interobserver concordance of echocardiographic assessment of right ventricular size and systolic function: a quality control exercise. J Am Soc Echocardiogr. 2012;25(7):709–713. doi:10.1016/j.echo.2012.03.018

32. Orde S, Slama M, Yastrebov K, Mclean A, Huang S; College of Intensive Care Medicine of Australia and New Zealand [CICM] Ultrasound Special Interest Group [USIG]. Subjective right ventricle assessment by echo qualified intensive care specialists: assessing agreement with objective measures. Crit Care. 2019;23(1):70. Published 2019 Mar 7. doi:10.1186/s13054-019-2375-z

33. Moceri P, Duchateau N, Sartre B, et al. Value of 3D right ventricular function over 2D assessment in acute pulmonary embolism. Echocardiography. 2021;38(10):1694–1701. doi:10.1111/echo.15167

34. Nagata Y, Wu VC, Kado Y, et al. Prognostic Value of Right Ventricular Ejection Fraction Assessed by Transthoracic 3D Echocardiography. Circ Cardiovasc Imaging. 2017;10(2):e005384. doi:10.1161/CIRCIMAGING.116.005384

35. Zhang Y, Sun W, Wu C, et al. Prognostic Value of Right Ventricular Ejection Fraction Assessed by 3D Echocardiography in COVID-19 Patients. Front Cardiovasc Med. 2021;8:641088. Published 2021 Feb 9. doi:10.3389/fcvm.2021.641088

36. Nambiar L, Volodarskiy A, Tak KA, et al. Acute COVID-19-Associated Decrements in Left and Right Ventricular Function Predict All-Cause Mortality. J Am Soc Echocardiogr. 2022;35(2):229–234. doi:10.1016/j.echo.2021.10.002

